# Unifying the communities of early-onset glycogen storage disease type IV and adult polyglucosan body disease through a genetic prevalence study of *GBE1*-related disease

**DOI:** 10.64898/2025.12.16.25342386

**Authors:** Rebecca L. Koch, H. Orhan Akman, Erin Chown, Deberah Goldman, Jeff Levenson, Qing Lu, Lindsay T. Michalovicz Gill, Matthew Morgan, Jennifer L. Orthmann-Murphy, Natacha T. Pires, Rebecca Reef, Harriet Saxe, Moriel Singer-Berk, Samantha Baxter

## Abstract

Glycogen storage disease type IV (GSD IV) is an autosomal recessive disorder caused by pathogenic variants in *GBE1*, resulting in deficient glycogen branching enzyme (GBE) activity and formation of abnormal glycogen (“polyglucosan”). GSD IV manifests across a spectrum of clinical dimensions – including hepatic, neurologic, muscular, and cardiac involvement – which vary in severity. The early-onset forms, historically referred to as Andersen disease, present at different stages ranging from in utero to adolescence. The adult-onset form, referred to as adult polyglucosan body disease (APBD), typically presents in middle to late adulthood. To date, no epidemiological study of GSD IV has been performed. Understanding the global prevalence of GSD IV is critical to increase disease awareness, improve diagnostic rates, inform therapeutic development, and engage pharmaceutical companies. In collaboration with the Rare Genomes Project at the Broad Institute of MIT and Harvard and the APBD Research Foundation, this study curated variants in *GBE1* and calculated prevalence across nine genetic ancestry groups. The estimated global carrier frequency of GSD IV is 1 in 243 individuals, and the global genetic prevalence is 1 in 235,784 individuals. Based on the 2024 world population, the estimated number of affected individuals with GSD IV is approximately 34,800. These estimates highlight a significant underdiagnosis of GSD IV and underscore the urgent need for increased awareness of this metabolic disorder. This model of collaboration between researchers, patient advocacy organizations, and genetic data sharing programs provides a framework for estimating the prevalence of other rare diseases in the global population.

**Graphical abstract:** Created in BioRender. Koch, R. (2025) https://BioRender.com/j0sg30n.

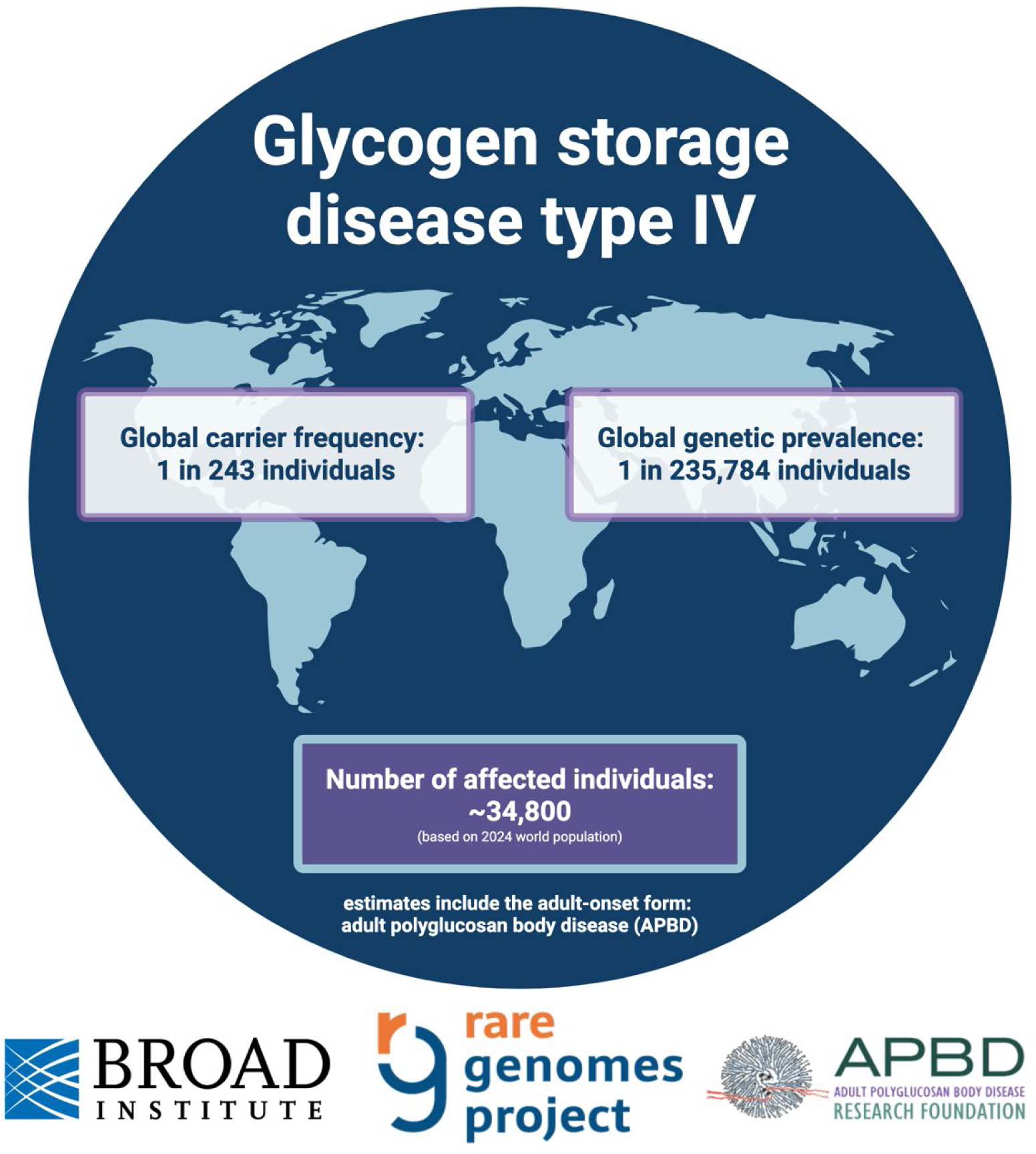

## INTRODUCTION

Glycogen storage disease type IV (GSD IV) is caused by biallelic pathogenic variants in *GBE1* (HGNC:4180) which results in reduced activity of glycogen branching enzyme (GBE, EC 2.4.1.18). As a result, glycogen synthesis is impaired and poorly branched, abnormal glycogen (“polyglucosan”) accumulates in cells, ultimately aggregating into polyglucosan bodies that disrupt cellular function [1]. Phenotypically, GSD IV is incredibly heterogeneous and is conceptualized as a multidimensional clinical continuum with hepatic, neurologic, muscular, and cardiac involvement occurring to varying degrees of severity [2]. The early-onset forms – initially referred to as Andersen disease and later classified as GSD IV (OMIM #232500) – include presentations in utero, during infancy, childhood, and adolescence. The adult-onset form, referred to as adult polyglucosan body disease (APBD, OMIM #263570), is a neurodegenerative disease with typical presentations in middle to late adulthood. Numerous pathogenic variants in *GBE1* have been reported with varying impacts on residual GBE activity levels, underscoring the disease heterogeneity [1]. However, the genotype-phenotype relationship remains poorly understood.

GSD IV, including the early-onset and APBD forms, is classified as an ultra-rare autosomal recessive disease and has been reported in various ethnic groups [2, 3]. Prevalence of a disease is an important factor in decision-making by researchers, pharmaceutical companies, and policymakers to determine goals and resource allocation toward the disease, yet the prevalence of the vast majority of rare diseases is unknown. Increased knowledge about the prevalence of a disease, both globally and in specific genetic ancestry groups, is important to patient advocacy and research organizations that aim to extend outreach and support to build a network that engages as many affected individuals and families as possible.

To date, a formal epidemiological study of all GSD IV phenotypes has not been performed. A general prevalence estimate of 1 in 600,000 to 800,000 individuals was proposed [4]. Since then, prevalence and carrier frequency studies have been focused on the APBD form. In 2012, the first epidemiological study of the *GBE1* c.986A>C (p.Tyr329Ser) variant was published [5]. This variant causes APBD when homozygous or compound heterozygous with a deep intronic variant [6]. It occurs at a higher frequency in the Ashkenazi Jewish ancestry group [3, 5], with this study estimating a carrier frequency of APBD of approximately 1 in 35 individuals of Ashkenazi Jewish ancestry [5]. Then, based on next-generation carrier screening performed in 2016, Sema4 expanded on this and included the c.986A>C (p.Tyr329Ser) variant along with other *GBE1* variants observed in patients with APBD; they estimated the carrier frequency for APBD in individuals of Ashkenazi Jewish ancestry to be 1 in 48 [7, 8]. Because the methods and results differed between the two studies, a follow-up investigation was performed in 2020 and estimated the prevalence of APBD in individuals of Ashkenazi Jewish ancestry over the age of 50 years old in the United States to be 3,400 to 6,400 [8]. Notably, this report also highlighted the high misdiagnosis rate of APBD with multiple sclerosis in the United States.

Despite the importance of these initial carrier frequency and prevalence studies on APBD, they are inherently limited in their application to the broader GSD IV community. The aforementioned studies did not include prevalence estimates of ethnicities other than Ashkenazi Jewish ancestry and were restricted to those with the APBD form rather than all patients with GSD IV. This study expands on previous efforts and produces estimates of the global carrier frequency and genetic prevalence of all GSD IV forms, including the early-onset and APBD forms. Appreciating the unmet medical need in the GSD IV community – which drives research into disease diagnosis, management, and treatment – requires that we develop a clearer understanding of the global prevalence of this disease.

## METHODS

### Nomenclature

Consistent with the clinical practice guidelines for GSD IV [1], we herein considered GSD IV to be a continuum of disease, varying by age of onset and organ involvement. In recognition of the APBD form having a relatively distinct collection of neurological symptoms and typical age of onset in adulthood, we refer to those with onset in childhood or adolescence as “early-onset GSD IV” and those with adult-onset GSD IV as “adult polyglucosan body disease” (APBD).

### Study design

In collaboration with the Rare Genomes Project at the Broad Institute of MIT and Harvard and the APBD Research Foundation, this study was conducted to estimate the global genetic prevalence of all known disease-causing variants in *GBE1* (GRCh37, ENST00000429644.2). A comprehensive list of suspected pathogenic variants in *GBE1* was assembled for variant curation according to American College of Medical Genetics and Genomics/Association for Molecular Pathology (ACMG/AMP) Guidelines for Sequence Variant Interpretation [9–11]. Sources for genetic variants included ClinVar (pathogenic/likely pathogenic/conflicting with at least one source saying pathogenic/likely pathogenic) [12], the Human Gene Mutation Database (HGMD; disease-causing mutation) [13], and a gene- and/or disease-specific database (https://digitalinsights.qiagen.com/), as well as any other high-confidence predicted loss-of-function (pLoF) variants in Genome Aggregation Database (gnomAD) [14, 15]. The duration of this project spanned two versions of gnomAD (v2 and v4). All variants that met our criteria in v2 received a full ACMG and pLoF curation, however due to the large number of new variants introduced in v4 (n = 200), we designed a triaged approach to review all new v4 variants. Any variant with an allele count (AC) ≥15 (allele frequency (AF) >0.00001) received a full ACMG curation, and any pLoF variant with AC>1 received LoF curation [14]. All variants which met our criteria in v2 but were initially classified as variants of uncertain significance (VUSs) were re-curated to ensure there was no new evidence that would change the classification.

Variants classified as VUSs from ClinVar and other databases were not included in this analysis as including these would inaccurately inflate the estimated prevalence of disease due to the potential of them being classified as benign when more evidence is gathered over time. In addition, the variants that were originally listed as pathogenic, likely pathogenic, or conflicting in ClinVar or disease-causing mutation in HGMD, but were classified as VUSs after curation, are included in this analysis as VUSs. Full details on the variant list creation and curation are found in the **Supplementary File**. After curation was performed, the allele frequencies were collected from gnomAD v4.1 and the aggregate carrier frequency was estimated on a global level, by distinct genetic ancestry groups. The Hardy-Weinberg principle was then applied where q = variant AF, Σq = aggregate AF, 2Σq = cumulative carrier frequency, (Σq)^2^ = genetic prevalence. “Conservative” prevalence estimates include only pathogenic and likely pathogenic variants (excluding VUSs), and “relaxed” prevalence estimates include pathogenic variants, likely pathogenic variants, and VUSs. The curated annotations of predicted protein-truncating variants were released to the public through the gnomAD browser, and the variant curations were submitted to ClinVar.

## RESULTS

Variant databases were queried to compile a list of variants thought to be associated with GSD IV, including early-onset and adult-onset phenotypes, in order to estimate the genetic prevalence for this recessive disease **(Figure 1)**. Each variant was standardized using the GRCh38 genome build and the canonical transcript NM_000158.4. After aggregating the list of variants from ClinVar, HGMD, and other gene- and disease-specific databases and accounting for duplicates, 104 variants met our criteria for full ACMG/AMP curation, and/or LoF curation [14], and 166 received an abbreviated curation (e.g., literature search). Ultimately, 4 variants were removed from consideration because they were classified as benign/likely benign. In the end, a total of 266 variants in *GBE1* were included in the carrier frequency and prevalence calculations: 8 pathogenic, 230 likely pathogenic, and 28 VUSs. Of note, the 28 VUSs do not include all VUSs from ClinVar, but rather the pathogenic/likely pathogenic variants that were downgraded to a VUS after a full ACMG/AMP curation.

**Figure 1.**
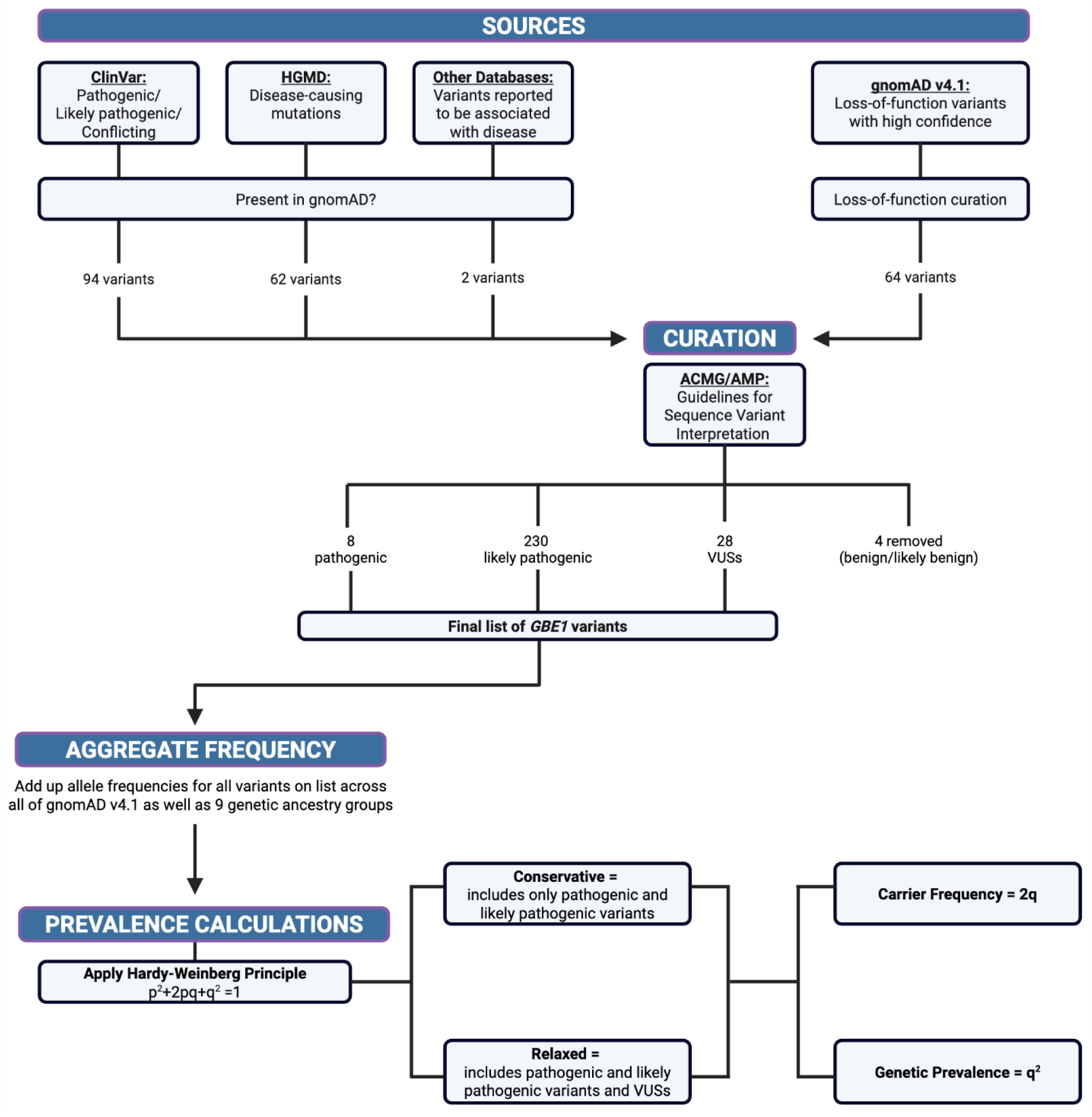
Overview of genetic prevalence study of GSD IV conducted by the Rare Genomes Project at the Broad Institute of MIT and Harvard in collaboration with the APBD Research Foundation. Created in BioRender. Koch, R. (2025) https://BioRender.com/7uj18lq.

Based on calculations with only pathogenic and likely pathogenic variants in *GBE1*, the estimated conservative global carrier frequency of all GSD IV phenotypes (i.e., early-onset and adult-onset) is 1 in 243 individuals, and the conservative global genetic prevalence of GSD IV is approximately 1 in 235,800 individuals **(Table 1**, **Figure 2)**. Based on the 2024 world population (8.2 billion), the conservative estimated number of affected individuals with GSD IV is approximately 34,800. The carrier frequency varied amongst genetic ancestry groups. As expected, there was a significantly higher conservative carrier frequency in the Ashkenazi Jewish group (1 in 64 individuals). The next most frequent carrier was the non-Finnish European group (1 in 227 individuals), followed by the East Asian group (1 in 265 individuals) and the Admixed American group (1 in 293 individuals). Based on the 2024 United States and European population estimates of 336 and 745 million, respectively, and the overall (all groups) carrier frequency, there are an estimated 1,452 individuals with GSD IV in the United States compared to an estimated 3,160 individuals with GSD IV in Europe. When specifying these estimates based on non-Finnish European carrier frequency, there are an estimated 3,625 affected individuals in Europe.

**Figure 2.**
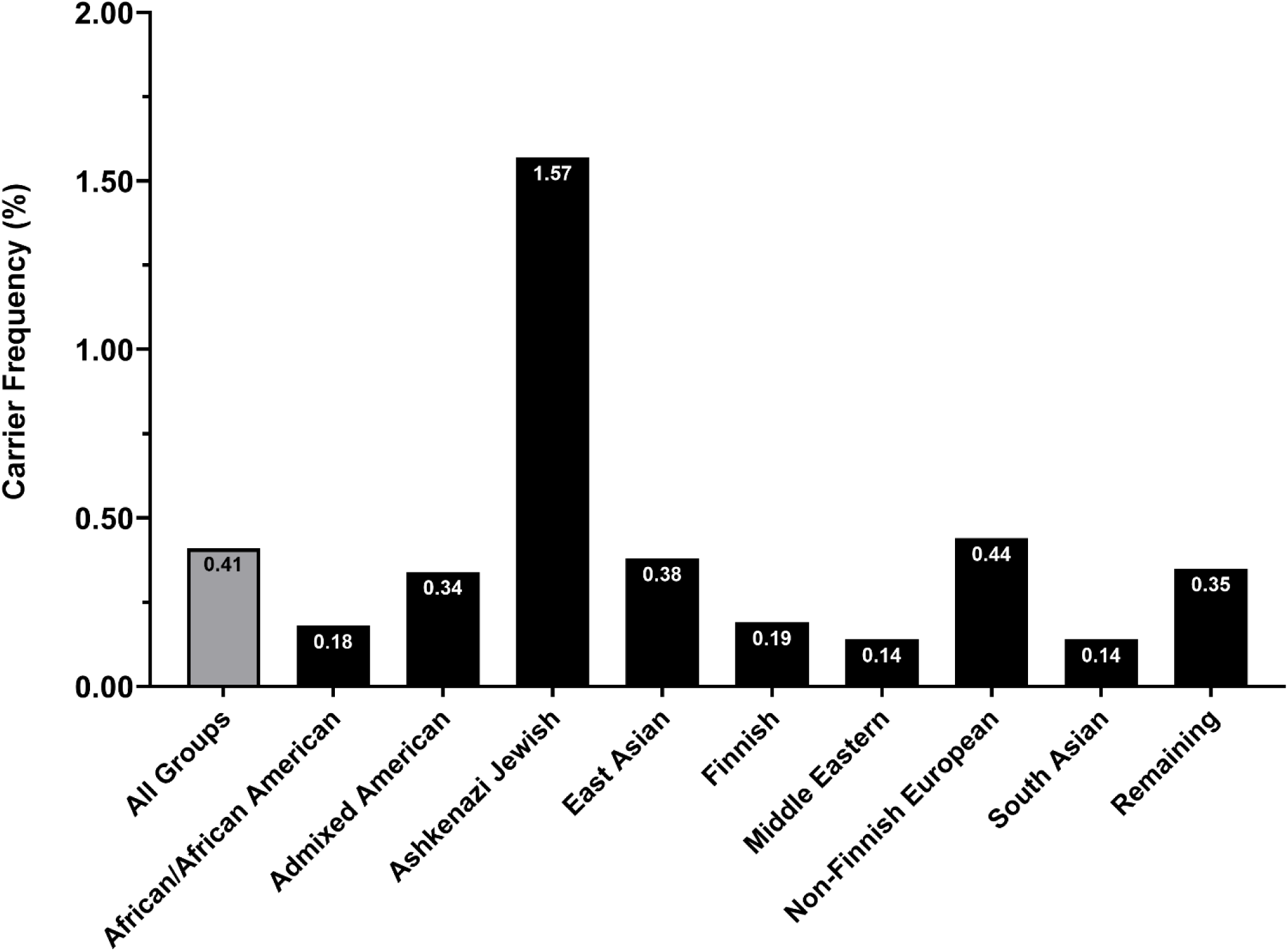
Group-specific carrier frequencies of GSD IV. The conservative carrier frequencies were calculated using pathogenic/likely pathogenic variants in *GBE1*. The carrier frequency (represented as a percentage of people) was rounded to the nearest hundredths value for individuals in all genetic ancestry groups (All Groups, gray) as well as by specific genetic ancestry groups (black).

**Table 1.** Global carrier frequency (2q) and genetic prevalence (q^2^) of GSD IV by genetic ancestry group. Calculations are based on conservative estimates which include pathogenic and likely pathogenic variants and exclude variants of uncertain significance.

|  | **# of**<br>**individuals** | **# of pathogenic/likely**<br>**pathogenic variants** | **Carrier**<br>**frequency** | **Genetic**<br>**prevalence** |
| --- | --- | --- | --- | --- |
| **All Groups** | 807,162 | 238 | 1/243 | 1/235,776 |
| **African/African American** | 30,019 | 32 | 1/564 | 1/1,270,243 |
| **Admixed American** | 37,545 | 32 | 1/293 | 1/342,705 |
| **Ashkenazi Jewish** | 14,804 | 5 | 1/64 | 1/16,258 |
| **East Asian** | 22,448 | 20 | 1/265 | 1/281,930 |
| **Finnish** | 32,026 | 11 | 1/531 | 1/1,127,547 |
| **Middle Eastern** | 3,031 | 4 | 1/733 | 1/2,148,714 |
| **Non-Finnish European** | 590,031 | 170 | 1/227 | 1/205,539 |
| **South Asian** | 45,546 | 31 | 1/738 | 1/2,180,633 |
| **Remaining** | 31,256 | 32 | 1/289 | 1/335,134 |

Among the 238 pathogenic and likely pathogenic variants in *GBE1*, the variants that occurred in the highest frequency across all genetic ancestry groups were c.691+2T>C (p.?, 0.20% carrier frequency) and c.986A>C (p.Tyr329Ser; 0.03% carrier frequency). The 691+2T>C variant was observed across all genetic ancestry groups except within the Admixed American, East Asian, and South Asian groups, with the highest incidence in the Non-Finnish European group (0.25% carrier frequency). The c.986A>C (p.Tyr329Ser) variant was most commonly observed in the Ashkenazi Jewish group (1.41% carrier frequency), and it was also observed – albeit more rarely – in the Non-Finnish European and the Remaining groups (0.006% and 0.10% carrier frequency, respectively). Though gene deletions and deep intronic variants have been reported in *GBE1* [16], whole genome sequencing data was only available from a select number of individuals in gnomAD v4 (76,215/807,162 total individuals [17]), and we were therefore not able to calculate their carrier frequencies.

The relaxed (i.e., including VUSs in *GBE1*) carrier frequency and genetic prevalence results are included in the **Supplementary File**.

## DISCUSSION

Understanding the carrier frequency and genetic prevalence of GSD IV is critical to improving diagnoses, supporting outreach to carriers and individuals with GSD IV, informing therapeutic development, and engaging with pharmaceutical companies.

This study builds on previous work by offering global prevalence estimates for all known pathogenic/likely pathogenic *GBE1* variants, capturing the full spectrum of GSD IV, from early-onset to adult-onset APBD. Pooling variants from ClinVar, HGMD, and gnomAD and classifying them according to the ACMG/AMP criteria, we calculated the genetic prevalence of GSD IV to be approximately 1 in 235,800 individuals – an estimate which includes individuals who are symptomatic, presymptomatic, as well as nonviable pregnancies. Based on the 2024 world population, there is an estimated 34,800 affected individuals with GSD IV. The large discrepancy between these calculations and the number of cases reported in the literature may be due to a high rate of misdiagnosis and delayed diagnosis. These persistent diagnostic challenges were highlighted during the APBD Patient-Led Listening Session with the FDA in October 2021 [18]; testimonies at the session revealed that patients often experience a constellation of symptoms for years, with reported diagnostic journeys that far exceed the cited average of 6.8 years [19], frequently misattributed to more common conditions (e.g., multiple sclerosis). Contributing factors were highlighted in the recent clinical practice guidelines for GSD IV and may include misdiagnoses, undergoing genetic testing with a targeted gene panel that excludes *GBE1*, failure to pursue further evaluation after negative or inconclusive results, and tests that do not cover deep intronic variants or large deletions [1]. Compounding these issues is a broader lack of awareness of GSD IV and its phenotypic continuum within the healthcare community. This study reinforces the urgent need to confront these barriers through improved diagnostic strategies, and continued advocacy for research and treatment for the GSD IV community.

The number of pathogenic *GBE1* variants and broad phenotypic heterogeneity of GSD IV depicts a complex relationship between genotype and phenotype. Interestingly, there have been reports of individuals with early-onset GSD IV or APBD with the same *GBE1* genetic variants [20, 21], patients with traditional features of hepatic early-onset GSD IV who later developed symptoms consistent with APBD in adulthood [21, 22], as well as patients who developed symptoms consistent with APBD but at an earlier age [23, 24]. Therefore, recent studies have suggested that GSD IV is a phenotypic continuum with a spectrum of involvement from neurologic, muscular, hepatic, and cardiac systems with varying degrees of severity [2]. Given this genotypic overlap [25], we were unable to calculate the genetic prevalence of the early-onset vs APBD forms. However, we were able to estimate carrier frequency and genetic prevalence estimates by genetic ancestry groups.

Early-onset GSD IV cases have been reported in Europe, the Middle East, South Asia, and North and South America, among others [2, 20]. Based on our genetic prevalence estimates, there are more affected individuals in Europe (3,160 individuals) than in the United States (1,452 individuals). In recent years, the APBD Research Foundation has taken steps to strengthen engagement within the European community [26]. This outreach is an essential step towards closing the gap between the global prevalence estimates and confirmed diagnoses, especially as improved awareness and diagnostic access are key to identifying misdiagnosed and undiagnosed individuals. Interestingly, despite previous reports of GSD IV in India and other South Asian populations [20], the carrier frequency was the lowest in this group at 1 in 738 individuals; this is likely explained by a reduced representation of these populations in gnomAD v4 [27], and the carrier frequency may actually be higher in certain ancestry groups, such as the Indian population. APBD has also been reported in several populations [24, 28–38], but is known to be more common in individuals of Ashkenazi Jewish genetic ancestry [25]. According to our calculations, the carrier frequency of GSD IV in Ashkenazi Jews is 1 in 64 individuals, slightly lower than previously reported estimates (1 in 35-48 individuals) [5, 7, 8]. It is important to note that our calculations include both early-onset GSD IV and APBD, whereas the previous estimates were specifically calculated for individuals with the APBD phenotype based on *GBE1* genotype, age of onset, or both. Moreover, our estimates were not able to include the c.2053-3358_2053-3350delins deep intronic variant associated with APBD [6]. Therefore, the previously reported estimates likely hold merit when estimating prevalence and carrier frequency specifically for “classic” APBD (i.e., homozygous for the c.986A>C (p.Tyr329Ser) variant in *GBE1* or compound heterozygous with the c.986A>C (p.Tyr329Ser) and the c.2053-3358_2053-3350delins variants [6]).

As emerging evidence suggests that certain variants in *GBE1* can lead to both early-onset GSD IV and APBD, there may be a growing rationale for broader inclusion of *GBE1* in reproductive/carrier screening panels for populations with a higher carrier frequency. The carrier frequency in the Ashkenazi Jewish genetic ancestry group (1 in 64) is comparable to that of other conditions routinely screened for in this population, falling somewhere between Canavan disease (1 in 40-82 individuals [39]) and Bloom syndrome (1 in 111-157 individuals [40]). Additionally, there is a known prevalence of GSD IV in the Mennonite communities [41] which was not captured in this dataset. Including *GBE1* on carrier screening panels may be a helpful solution to the pervasive problem of misdiagnosis and may help close the gap between predicted and currently identified affected individuals. However, the ethical implications of screening for a disease with an unpredictable age of onset – from childhood to adulthood – as well as lack of disease-modifying therapies warrant further exploration and discussion, as most of these panels test for pediatric-onset conditions. A similar concern has been raised for carrier screening panels that test for Gaucher disease given carriers of pathogenic variants in *GBA1* are at a higher risk of developing Parkinson’s disease compared to the general population [42].

To date, there are no treatment options for GSD IV, and care remains focused on managing symptoms. Organ transplantation of the liver or heart is recommended for patients with liver or heart failure, respectively; yet, this strategy will not address the extra-hepatic and extra-cardiac disease manifestations [1]. However, a range of promising therapeutic approaches are under investigation **(Figure 3).** To understand these strategies, it helps to look deeper at glycogen synthesis. Glycogen is synthesized through the concerted actions of two main enzymes: i) glycogen synthase (GYS, EC 2.4.1.11), which adds glucose molecules to the end of growing linear chains, and ii) GBE, which transfers short pieces of these chains to create branches in the glycogen molecule. This highly branched structure is understood to be important for glycogen solubility and proper metabolism [43, 44]. Deficient GBE activity in GSD IV results in the formation of poorly branched, insoluble glycogen (“polyglucosan”) which disrupts cellular function and leads to disease symptoms [1]. In theory, dietary interventions with adjustment of carbohydrate intake could reduce glucose levels and thus there would be less substrate to form polyglucosan [1, 45]; however, this is of theoretical benefit and no prospective investigations have been conducted. Similarly, sodium-glucose cotransporter 2 (SGLT2) inhibitors have been proposed for therapeutic use in Lafora disease – a related GSD with polyglucosan accumulation – due to their pharmacological blockade of glucose reabsorption in the kidneys and subsequent excretion of glucose in the urine [46, 47], yet they have not been formally investigated in GSD IV. Researchers have also investigated adeno-associated virus (AAV)-mediated downregulation or knockout of *GYS1* (the GYS isoform expressed in most tissues) to reduce total GYS1 activity [48, 49]. There are GYS1-targeting therapies being investigated for other GSDs which theoretically hold promise for GSD IV, such as intrathecally administered ASO therapy targeting GYS1 for Lafora disease (ION283, NCT06609889), or RNA interference-mediated GYS1 reduction for Pompe disease [50] (ABX1100, NCT06109948). In addition, gene replacement therapy candidates currently under investigation aim to restore GBE activity [51]. Meanwhile, n-Lorem is pursuing a precision antisense oligonucleotide (ASO) therapy for a small group of APBD patients with a deep intronic variant in *GBE1* [6, 52]. Using mRNA therapy to restore GBE activity levels has not been investigated in GSD IV but is of theoretical benefit.

**Figure 3.**
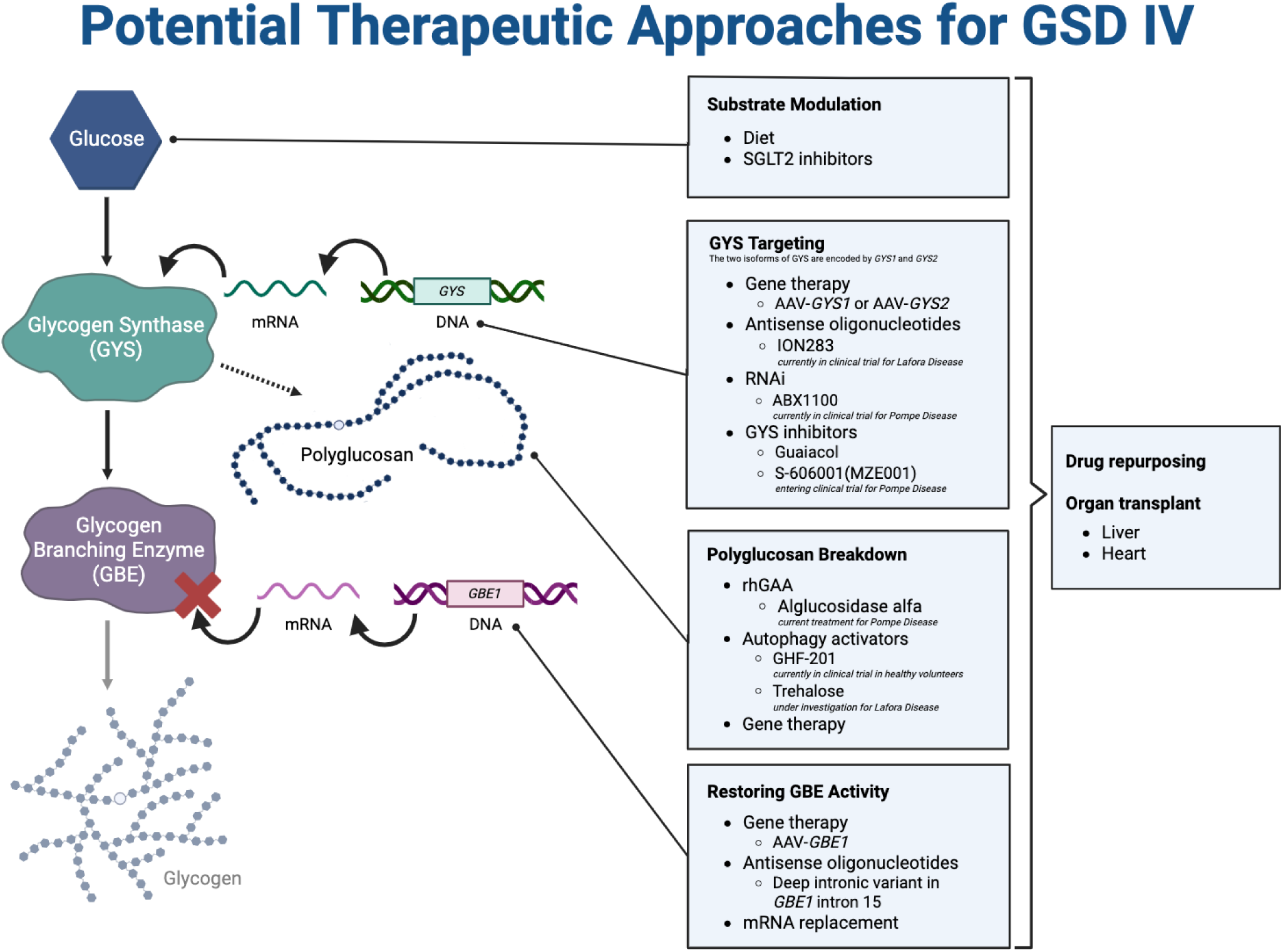
Prospective therapeutic approaches for GSD IV. There is a range of therapeutic approaches under investigation for GSD IV and related GSDs, such as Lafora disease and Pompe disease. Many of these approaches target different aspects of the glycogen synthesis (left) and polyglucosan breakdown. These approaches include substrate modulation, reducing total glycogen synthase (GYS) activity (GYS1 or GYS2 isoforms), breaking down accumulated polyglucosan, and restoring glycogen branching enzyme (GBE) activity. This list is not exhaustive, rather it offers potential therapeutic opportunities that may or may not have been investigated. Additional abbreviations: AAV, adeno-associated virus; DNA, deoxyribonucleic acid; mRNA, messenger ribonucleic acid; rhGAA, recombinant human acid alpha-glucosidase; RNAi, ribonucleic acid interference. Created in BioRender. Gill, L. (2025) https://BioRender.com/6v460xc.

In addition to these gene- and RNA-based interventions, researchers are also investigating treatments that work through alternative mechanisms. One promising small molecule candidate is GHF-201 (compound 144DG11, Kakhlon et al., 2021), a small molecule developed by Golden Heart Flower Ltd. that is currently being assessed in healthy adult volunteers (EUCT 2024-519813-74-00). Supplementation of guaiacol (a GYS inhibitor) [53] and treatment with alglucosidase alfa (to enhance lysosomal polyglucosan degradation) [54] have been explored as potential therapeutics to reduce polyglucosan accumulation in GSD IV. Compounds initially tested in other related diseases may be worth investigating in GSD IV, such as trehalose – an inducer of autophagy proposed as a therapeutic in Lafora disease [55, 56] – or S-606001 (formerly MZE001) – a small molecule and specific inhibitor of GYS1 being investigated as a potential oral treatment for Pompe disease (NCT07123155) [57]. It is important to note that this list is not exhaustive and with the improvements in technologies like machine learning models, there may be opportunities to repurpose drugs for the treatment of GSD IV [58].

As the therapeutic pipeline for GSD IV continues to grow, genetic prevalence studies become increasingly critical to aid in identifying how many individuals are affected by the disease. Importantly, prevalence estimates can inform the design of clinical trials, support regulatory planning, and guide investment in therapeutic development, ensuring that emerging treatments reach the patient populations for whom there currently are no approved treatment options.

This study estimated carrier frequency and disease prevalence for recessive conditions using curated variants in *GBE1*. This methodology has been used for genetic prevalence studies of several genetic diseases [59], such as for phospholipase A2 group VI (PLA2G6)-associated neurodegeneration which was done by the Rare Genomes Project at the Broad Institute of MIT and Harvard [60]. There are many factors that go into calculating the estimated carrier frequencies and genetic disease prevalence, such as available data in ClinVar (database of classified variants) and gnomAD (reference population database), as well as curation guidelines. It is important to consider that these calculations are estimates and include inherent limitations. Individuals with rare diseases are less likely to participate in or meet recruitment criteria for research studies that are included in reference population databases, so this method is only appropriate for autosomal recessive diseases and some X-linked recessive diseases, not X-linked or autosomal dominant conditions. For the same reason, the depletion of symptomatic carriers could lower the allele frequencies of pathogenic variants observed in gnomAD. Moreover, there are some reported pathogenic and likely pathogenic variants that were not found in gnomAD and therefore could not be included in this analysis. While gnomAD is the world’s largest human reference database, it is not representative of the entire global population. Some genetic ancestry groups are either underrepresented or still missing from gnomAD and more diversity of reference data is needed. We expect future releases of gnomAD to increase representation of diverse genetic ancestries; however, this is an area that will continue to need to be addressed. Future versions of gnomAD are expected to include more samples with whole genome sequencing which will improve calculation estimates of variants that are not detected with whole exome sequencing, such as deep intronic variants and large deletions. Finally, this calculation does not take into account the increased rates of consanguinity in some cultures which increase the prevalence of recessive conditions in these regions. Lastly, we excluded VUSs – which were often missense variants – from our conservative estimates, yet with GBE testing to biochemically support the diagnosis as well as improved understanding of the effect of these missense variants on the protein function, it is expected that many VUSs may be able to be reclassified as pathogenic or likely pathogenic. It is important to note that the prevalence numbers reported here may change in the future as technologies evolve and allele frequencies are refined with more uniform data sharing policies. Moreover, these prevalence estimates may change as natural history data is published and our understanding of the genotype-phenotype relationships improves.

## CONCLUSION

Our genetic prevalence study of GSD IV estimated that there are approximately 34,800 individuals affected by GSD IV globally, with an overall carrier rate of 1 in 243 individuals. Though the APBD form remains prevalent in the Ashkenazi Jewish group, this study highlights the genetic prevalence of GSD IV across a variety of ancestry groups. This comprehensive genetic prevalence study paves the way for future clinical trial design as researchers assess the potential impact of therapies for GSD IV on the global population. In addition, having a more accurate prevalence estimate is essential for reducing the rate of misdiagnosis and diagnostic delay that affected patients often report experiencing. This model of collaboration between researchers, patient advocacy organizations, and genetic data sharing programs provides a framework for estimating the prevalence of other rare diseases in the global population.

## Data Availability

All estimated genetic prevalence reports are publicly available through the Genetic Prevalence Estimator (GeniE) tool. Specifically, the conservative estimates (i.e., only includes pathogenic and likely pathogenic variants in GBE1) are available at https://broad.io/GeniE_GBE1 and the relaxed estimates (i.e., includes pathogenic and likely pathogenic variants as well as variants of uncertain significance in GBE1) are available at https://broad.io/GeniE_GBE1_relaxed.

https://broad.io/GeniE_GBE1_relaxed

## ACKNOWLEDGEMENTS

The authors gratefully acknowledge the Kathryn Russell, Carmen Glaze, Jordan Wood, Emily Evangelista, Anne O’Donnell-Luria, and Heidi Rehm for their support and analysis, and the Y.T. and Alice Chen Pediatric Genetics and Genomics Research Center for its encouragement of this work. The authors would like to also thank the APBD Research Foundation and the GSD IV disease community for their support and helpful discussions, as well as the Keith B. Hayes Foundation for its support of glycogen storage disease research.

## Notes

### Competing Interest Statement

Rebecca L. Koch has received grant/funding support from Alnylam Pharmaceuticals. Rebecca L. Koch and H. Orhan Akman have received grant/funding support from the Keith B. Hayes Foundation. Jennifer Orthmann-Murphy is a consultant for Vigil Neuroscience and Savanna Bio as well as a site PI for Vigil Neuroscience clinical trials. Samantha Baxter is a paid consultant for Pharming Group N.V. The Adult Polyglucosan Body Disease Research Foundation is a shareholder of Golden Heart Flower, Ltd., holds a revenue sharing agreement with Columbia University for treatment development of guaiacol, and has received grant/funding support from the Chan Zuckerberg Initiative. All other authors declare no conflict of interest.

### Funding Statement

Parts of this study were funded by the Chan Zuckerberg Initiative Donor-Advised Fund at the Silicon Valley Community Foundation 2020-224274 and 2022-316726 (https://doi.org/10.37921/236582yuakxy) (funder DOI 10.13039/100014989).

### Author Declarations

The study used openly available human data that were originally located at: ClinVar, the Human Gene Mutation Database (HGMD), and Genome Aggregation Database (gnomAD).

