## Supplementary Material for "Unifying the communities of early-onset glycogen storage disease type IV and adult polyglucosan body disease through a genetic prevalence study of *GBE1*-related disease"

### METHODS SUMMARY

*Prepared by the Rare Genomes Project – Disease Prevalence Study Rare as One Network team*

#### Variant List Creation:

To compile the list of variants for ACMG curation, we collected all pathogenic and likely pathogenic variants from ClinVar, including any listed as conflicting where one or more submissions categorized the variant as pathogenic or likely pathogenic. Additionally, we collected all variants from the Human Gene Mutation Database (HGMD) categorized as "disease mutation (DM)"[1] and variants provided to us by the APBD Research Foundation. We then cross referenced these lists with variants that passed gnomAD v4.1 QC filters and were identified in at least one individual in gnomAD.[2]

For loss of function (LoF) curation, we collected the list of all high confidence variants from gnomAD that passed QC where the transcript consequences are in the set ("stop_gained", "frameshift_variant", "splice_acceptor_variant", "splice_donor_variant", "transcript_ablation"). When applicable, some variants received both ACMG curation and LoF curation.

#### Variant Curation:

*ACMG curation*: Variant classifications are based on ACMG/AMP criteria[3] with ClinGen rule specifications.[4] All sequence variants were curated using the ClinGen Variant Curation Interface[5] and were classified using the ACMG-recommended five-tier classification system: Pathogenic, Likely Pathogenic, Uncertain Significance, Likely Benign, Benign.

*Loss of Function Curation*: Loss-of-Function (LoF) curation consists of manual review of predicted LoF variants[6] for the presence of information that indicates the variant would not lead to LoF of the gene **(Supplementary Table 1)**. These error modes include: technical artifacts in read mapping, variant calling or QC, rescue events, and biological low importance. After reviewing each variant, the variant was scored using a five point scale: not LoF, likely not LoF, uncertain, likely LoF, and LoF. In order for a variant to be considered as LoF, it had to have no major error modes selected (such as a rescue event). Variants were classified as likely LoF if a single minor error mode was noted for a variant, which include some genotyping or mapping errors, or weak exon conservation/minority of transcripts with high exonic expression. In contrast, rescue events, for example, were automatically classified as likely not LoF or not LoF. Multiple error modes ($\geq$3) generally resulted in a "not LoF" curation of the variant. Variants in which there was inconclusive evidence supporting the variant as LoF or not LoF were curated as uncertain. Each variant was independently curated by two curators before being assessed to eliminate disagreements. A literature search was performed on each of these variants, and all variants were classified using the ACMG-recommended five-tier classification system: Pathogenic, Likely Pathogenic, Uncertain Significance, Likely Benign, Benign.

**Supplementary Table 1. Loss-of-Function (LoF) curation and error mode descriptions.**

| LoF Error Code | Description |
|---|---|
| Mapping Error | There may be issues with mapping of reads to this region that overlaps the variant. |
| Genotyping Error | Quality of the reads for individuals with the variant is low, particularly for read depth, allele balance, and/or genotype quality. |
| Homopolymer | Variant is an insertion/deletion that is found within a homopolymer repeat (>=5 repeats). |
| No Read Data | Individual reads from gnomAD are not available for viewing for individuals with this variant because of data access issues. This can happen when the raw sequence data is no longer available to the gnomAD team at the time of generating these images. |
| Reference Error | Variant is due to an error with the human reference genome build and is not real. |
| Strand Bias | Reads of individuals with the variant show a strong bias for either the forward or reverse strand. |
| MNV/Frame-restoring indel | MNV (multi-nucleotide variant) in which there are two variants that are occurring in the same codon. One of these alone would cause a nonsense variant, but both together and in cis would only cause a synonymous or missense variation. Variants that are in cis with other indels that could restore the frame are called frame-restoring indels, and would result in an in-frame indel instead of loss of function. |
| Essential Splice Rescue | Variant that disrupts the +/-1,2 essential splice site, but in which there is an in-frame cryptic splice site that is predicted to rescue the transcript by spliceAI/pangolin. |
| In-frame exon | Variant that disrupted the +/-1,2 essential splice site, but there is a prediction of in-frame exon skipping by splicAI/pangolin that is predicted to rescue the transcript |
| Minority of Transcripts | Variant exists in a minority of the transcripts present for that gene. |
| Weak Exon Conservation | Variant falls within an exon that is weakly conserved relative to the rest of the gene. |
| Last Exon | Variant (or stop codon for a frameshift variant) falls within the last exon for the gene or the last 50 base pairs of the penultimate exon for the gene. |
| Other Transcript Error | Variant falls within a transcript that is overprinted (more than 1 transcript with different frames), is in a region of an exon that is overhanging other exons and weakly conserved in the overhanging regions, or is a nonsense variant within the first coding exon and in which there is a downstream Met that could reinitiate translation. |
| Low pext (<0.2) | Variant falls within an exon that has a low pext score relative to the highest score across the gene. The pext scores represent exon level expression from GTEx tissues. |
| Pext < 50% max | Variant falls within an exon that has a slightly reduced pext score relative to the highest score across the gene. The pext scores represent exon level expression from GTEx tissues. |

**Prevalence Calculation:**

Prevalence was calculated using the Hardy-Weinberg principle **(Supplementary Table 2).**

**Supplementary Table 2. Frequency calculation descriptions.**

| **Frequency Category** | **Definition** |
|---|---|
| Conservative Aggregate Carrier Frequency (2q) | This is the carrier frequency for the designated population for only variants that were classified as Pathogenic (P) or Likely Pathogenic (LP) using ACMG criteria for variant curation. Calculated by timesing the aggregated allele frequency of all LP/P variants by 2. |
| Conservative Prevalence ($q^2$) | This is the calculated prevalence for the disease in that given population, using aggregated allele frequency for Pathogenic (P) and Likely Pathogenic (LP) variants in that gene. Calculated by squaring the aggregated allele frequency (q) for P and LP variants in that gene. |
| Relaxed Aggregate Carrier Frequency (2q) | This is the carrier frequency for the designated population for only variants that were classified as Pathogenic (P), Likely Pathogenic (LP) and variants of uncertain significance (VUS) using ACMG criteria for variant curation. Calculated by timesing the aggregated allele frequency of all P/LP/VUS variants by 2. |
| Relaxed Prevalence ($q^2$) | This is the calculated prevalence for the disease in that given population, using aggregated allele frequency for Pathogenic (P) and Likely Pathogenic (LP) variants in that gene. Calculated by squaring the aggregated allele frequency (q) for P/LP/VUS variants in that gene. |

**Population and genetic ancestry group inference:**

Continental ancestry was inferred using a PCA and random forest approach. First and second degree relatives are removed from the dataset to have a dataset of unrelated individuals. Ancestries are assigned taking samples of known ancestry and then using the common genetic variants in the data to identify samples with genetic similarity. We do this by computing the top 20 principal components on the alternate allele counts for the same set of variants used in the PCRelate PCA and projecting the remaining related samples onto these principal components. Next, we trained a random forest model on a set of samples with known continental ancestry and used this model to assign continental ancestry labels to samples for which the random forest probability >0.9. For additional details on population inference, including how the subpopulations were determined, please see pages 13-16 of the supplementary materials in Karczewski et al. 2020.[7]

**RELAXED CALCULATION RESULTS (INCLUDING VUS IN *GBE1*)**

**Supplementary Table 3. Global carrier frequency (2q) and genetic prevalence ($q^2$) of GSD IV by genetic ancestry group.**

Calculations are based on relaxed estimates which include pathogenic and likely pathogenic variants as well as variants of uncertain significance (VUS).

| | # of individuals | # of pathogenic/likely pathogenic/VUS variants | Carrier frequency | Genetic prevalence |
|---|---|---|---|---|
| **All Groups** | 807,162 | 266 | 1/132 | 1/69,741 |
| **African/African American** | 30,019 | 39 | 1/409 | 1/667,594 |
| **Admixed American** | 37,545 | 39 | 1/131 | 1/68,527 |
| **Ashkenazi Jewish** | 14,804 | 7 | 1/61 | 1/14,692 |
| **East Asian** | 22,448 | 23 | 1/248 | 1/245,562 |
| **Finnish** | 32,026 | 15 | 1/255 | 1/260,114 |
| **Middle Eastern** | 3,031 | 7 | 1/41 | 1/6,604 |
| **Non-Finnish European** | 590,031 | 193 | 1/121 | 1/58,782 |
| **South Asian** | 45,546 | 39 | 1/224 | 1/200,459 |
| **Remaining** | 31,256 | 39 | 1/170 | 1/116,113 |

**Supplementary Figure 1. Group-specific carrier frequencies of GSD IV.**

The relaxed carrier frequencies were calculated using pathogenic/likely pathogenic variants as well as variants of uncertain significance (VUS) in *GBE1*. The carrier frequency percentage (%) was rounded to the nearest hundredths value for individuals in all populations (gray) as well as by subpopulations (black).

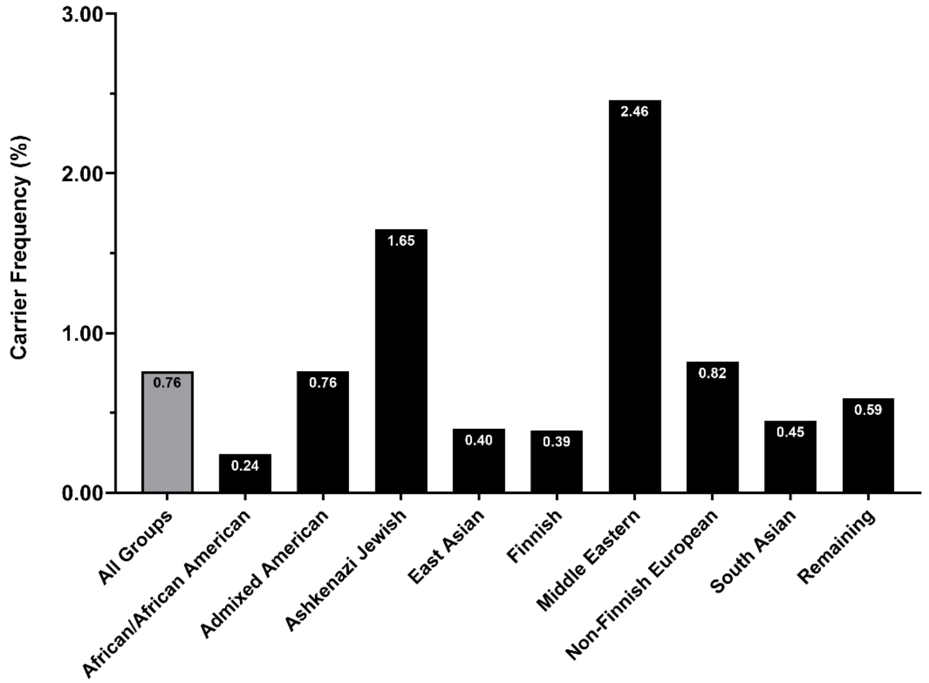
